# Triplex Real Time RT-PCR for N1, N2 and RP probes from CDC EUA SARS-CoV-2 diagnosis kit

**DOI:** 10.1101/2020.06.29.20133363

**Authors:** Byron Freire-Paspuel, Patricio Vega-Mariño, Alberto Velez, Marilyn Cruz, Miguel Angel Garcia-Bereguiain

**Affiliations:** One Health Research Group. Universidad de las Americas. Quito. Ecuador; Agencia de Regulación y Control de la Bioseguridad y Cuarentena para Galápagos. Puerto Ayora. Ecuador

## Abstract

CDC protocol for severe acute respiratory syndrome coronavirus 2 (SARS-CoV-2) include 3 targets for detection (N1, N2 and RP) labelled with FAM so 3 PCR reactions are required per sample. We developed a triplex, real-time reverse transcription PCR for SARS-CoV-2 that maintained clinical performance compared with CDC singleplex assay. This protocol could speed up detection and save reagents during current SARS-CoV-2 testing supplies shortage.

## Background

Severe acute respiratory syndrome coronavirus 2 (SARS-CoV-2) diagnosis relies on molecular detection of viral RNA from oropharyngeal or nasopharyngeal specimens. Multiple Real-time reverse transcription PCR (RT-qPCR) protocols have been developed for SARS-CoV-2 detection. Among them, the CDC protocol (1,2) is considered a gold standard worldwide as it was the first one endorsed by Emergency Use Authorization (EUA) by Federal Drug Administration (FDA) (3). The CDC designed 2019-nCoV CDC EUA kit (IDT, USA) is based on 3 singleplex RT-qPCR reactions for N1 and N2 probes to detect SARS-CoV-2, and and RNaseP (RP) as an RNA extraction quality control (1,2). This singleplex PCR protocol uses large amounts of reagents and reduce laboratory testing capacity, specially at small scale facilities, both of which have become crucial during the ongoing coronavirus disease pandemic, particularly at developing countries. Although several multiplex assays are commercially available, they rely on specific platforms or providers, and also are substantially more expensive.

## Objective

Our goal was to develop a triplex assay to detect SARS-CoV-2 RNA from nasopharyngeal swabs based on the CDC designed probes and primers N1, N2 and RP, and evaluate its performance using singleplex 2019-nCoV CDC EUA kit as a gold standard.

## Study design

70 clinical specimens (nasopharyngeal swabs) were included on this study. Samples were tested for SARS-CoV-2 following an adapted version of the CDC protocol (1,2) using AccuPrep Viral RNA extraction kit (Bioneer, South Korea), TaqMan Fast Virus 1-Step Master Mix (Applied Byosistems, USA) and CFX96 Thermocycler (BioRad). Triplex reactions contained each primer and probe in a final concentration of 0.5 and 0.13 µM, respectively.

N1 probe was labelled with FAM dye and BHQ1 quencher, N2 probe was labelled with HEX dye and BHQ1 quencher, and RP probe was labelled with Cy5 dye and TAO and Iowa Black RQ. All primers and probes used for the singleplex and triplex assay were purchased from IDT (USA) (see supplementary material).

## Results

Same RNA samples were tested for both single and triplex RT-qPCR assays. From the 70 samples included on the study, 50 tested positive and 20 tested negatives for SARS-CoV-2 using the singleplex 2019-nCoV CDC EUA kit (Table 1 and 2). All 20 negative samples for singleplex assay were also negative for the triplex assay. From the 50 positive samples by singleplex, 46 samples show amplification for N1 and N2 for the triplex assay, 3 samples show amplification for one viral probe (1 sample only N1; 2 samples only N2), and 1 sample show no viral probe amplification (Table 2). The Ct values obtained for N1 and N2 for singleplex assay were 30.70±3.83 and 31.34±4.09, respectively. The Ct values obtained for N1 and N2 for triplex assay were 30.24±3.82 and 32.57±4.69, respectively (no statistically significant differences for Ct values among both assays, p=0.559 and p=0.169).

**Table 1.** Performance of triplex RT-qPCR assay compared to singleplex 2019-nCoV CDC EUA (% value means sensitivity).

|  | Singleplex SARS-CoV-2 positive | Singleplex SARS-CoV-2 Negative |
| --- | --- | --- |
| **Triplex SARS CoV-2 Positive** | 49 (98%) | 0 |
| **Triplex SARS CoV-2 Negative** | 1 | 21 |

**Table 2.** Ct values and viral loads for triplex and singleplex nCoV-QS and 2019-nCoV CDC EUA RT-qPCR assays (samples highlighted: N1 positive or N1/N2 negative for triplex assay).

|  | Viral Load (c/μL) | N1 Ct Value | N2 Ct Value | RP Ct Value | N1-TX Ct Value | N2-TX Ct Value | RP-TX Ct Value |
| --- | --- | --- | --- | --- | --- | --- | --- |
| 1 | 386.000 | 21,22 | 21,98 | 25,13 | 20,25 | 22,41 | 25,15 |
| 2 | 255.000 | 21,27 | 21,34 | 23,25 | 20,82 | 22,56 | 24,01 |
| 3 | 45.900 | 23,41 | 24,03 | 25,01 | 23,76 | 25,29 | 25,75 |
| 4 | 22.300 | 24,48 | 24,08 | 30,08 | 23,96 | 24,78 | 31,48 |
| 5 | 15.100 | 25,05 | 24,7 | 27,45 | 24,96 | 25,76 | 28,18 |
| 6 | 13.500 | 24,88 | 24,92 | 25,53 | 24,59 | 25,78 | 26,58 |
| 7 | 7.740 | 25,7 | 25,65 | 25,45 | 25,37 | 27,31 | 25,18 |
| 8 | 5.940 | 26,72 | 27,74 | 25,14 | 25,95 | 28,27 | 25,26 |
| 9 | 4.360 | 26,54 | 26,66 | 26,36 | 26,58 | 28,08 | 27,54 |
| 10 | 3.090 | 28,47 | 29,43 | 33,52 | 28,2 | 30,02 | 33,69 |
| 11 | 2.920 | 27,48 | 27,64 | 28,9 | 27,71 | 28,38 | 29,63 |
| 12 | 2.060 | 27,65 | 27,47 | 28,07 | 27,13 | 28,32 | 29,1 |
| 13 | 1.870 | 29,33 | 30,18 | 25,35 | 29,03 | 31,46 | 26,45 |
| 14 | 1.540 | 29,65 | 30,37 | 27,01 | 29,64 | 31,05 | 27,84 |
| 15 | 1.270 | 28,77 | 30,18 | 25,23 | 29,03 | 30,72 | 25,97 |
| 16 | 956 | 29,13 | 29,56 | 27,59 | 29,29 | 31,05 | 28,79 |
| 17 | 763 | 29,11 | 29,47 | 28,89 | 28,62 | 30,02 | 29,47 |
| 18 | 693 | 29,73 | 31,34 | 24,36 | 29,92 | 32,32 | 25,46 |
| 19 | 690 | 31,03 | 31,65 | 22,6 | 31,21 | 33,63 | 23,62 |
| 20 | 612 | 29,72 | 30,05 | 25,15 | 28,56 | 31,13 | 25,59 |
| 21 | 509 | 30,21 | 31,03 | 27,02 | 30,15 | 31,88 | 27,59 |
| 22 | 306 | 30,06 | 31,68 | 19,57 | 30,65 | 33,39 | 22,15 |
| 23 | 271 | 30,99 | 31,4 | 31,4 | 30,33 | 31,37 | 32,34 |
| 24 | 189 | 31,52 | 31,85 | 30,25 | 31,14 | 32,99 | 30,99 |
| 25 | 175 | 31,63 | 31,99 | 28,2 | 31,46 | 32,47 | 29,54 |
| 26 | 164 | 31,36 | 31,49 | 31,58 | 30,5 | 31,88 | 32,73 |
| 27 | 160 | 31,13 | 31,55 | 23,57 | 31,79 | 34,45 | 25,37 |
| 28 | 152 | 31,48 | 32,32 | 23,99 | 32,28 | 35,2 | 25,04 |
| 29 | 139 | 31,98 | 33,06 | 24,63 | 32,56 | 34,44 | 26,33 |
| 30 | 123 | 32,15 | 33,79 | 31,3 | 32,31 | 34,57 | 32,52 |
| 31 | 102 | 32,08 | 32,58 | 28,79 | 30,88 | 33,52 | 29,1 |
| 32 | 870 | 32,66 | 32,87 | 32,72 | 33,06 | 34,02 | 33 |
| 33 | 69,9 | 32,24 | 33,31 | 24,92 | 32,98 | 35,3 | 26,13 |
| 34 | 69 | 32,59 | 33,33 | 25,56 | 31,68 | 33,98 | 25,98 |
| 35 | 60,9 | 32,82 | 34,46 | 33,77 | 32,27 | 34,00 | 33,42 |
| 36 | 59,3 | 33,23 | 33,62 | 27,05 | 34,25 | 34,87 | 27,86 |
| 37 | 54,6 | 33,35 | 34,33 | 27,1 | 34,18 | 35,93 | 28,07 |
| 38 | 43,9 | 32,95 | 37,54 | 28,62 | 34,19 | 37,44 | 30,59 |
| 39 | 33,6 | 34,07 | 33,98 | 30,28 | 34,1 | 40,31 | 32,15 |
| **40** | **33,4** | **34,08** | **36,89** | **25,1** | **35,1** | **N/A** | **27,01** |
| **41** | **22,4** | **34,67** | **35,63** | **25,16** | **N/A** | **36,39** | **26,24** |
| 42 | 19,7 | 33,93 | 32,54 | 25,52 | 32,99 | 32,73 | 25,42 |
| 43 | 16,4 | 34,17 | 35,41 | 23,61 | 33,88 | 41,25 | 24,44 |
| 44 | 12,7 | 35,51 | 37,97 | 29,42 | 35,69 | 42,54 | 30,75 |
| 45 | 12,6 | 34,53 | 33,69 | 27,43 | 33,16 | 36,33 | 29,22 |
| 46 | 11,9 | 35,21 | 35,65 | 24,08 | 34,77 | 40,14 | 24,86 |
| **47** | **10,3** | **35,42** | **34,25** | **35,3** | **N/A** | **35,18** | **35,79** |
| 48 | 9,25 | 34,94 | 35,24 | 27,48 | 34,37 | 37,53 | 29,36 |
| 49 | 3,20 | 37,14 | 37,16 | 26,21 | 35,95 | 40,9 | 27,05 |
| **50** | **1,56** | **37,33** | **37,94** | **24,39** | **N/A** | **N/A** | **24,73** |
| 51 | 0 | N/A | N/A | 34,18 | N/A | N/A | 36,07 |
| 52 | 0 | N/A | N/A | 30,66 | N/A | N/A | 31,9 |
| 53 | 0 | N/A | N/A | 24,18 | N/A | N/A | 24,67 |
| 54 | 0 | N/A | N/A | 27,16 | N/A | N/A | 27,15 |
| 55 | 0 | N/A | N/A | 23,38 | N/A | N/A | 24,1 |
| 56 | 0 | N/A | N/A | 24,63 | N/A | N/A | 25,46 |
| 57 | 0 | N/A | N/A | 24,96 | N/A | N/A | 25,69 |
| 58 | 0 | N/A | N/A | 23,05 | N/A | N/A | 24,3 |
| 59 | 0 | N/A | N/A | 20,66 | N/A | N/A | 21,74 |
| 60 | 0 | N/A | N/A | 23,53 | N/A | N/A | 24,44 |
| 61 | 0 | N/A | N/A | 22,76 | N/A | N/A | 23,31 |
| 62 | 0 | N/A | N/A | 21,88 | N/A | N/A | 22,37 |
| 63 | 0 | N/A | N/A | 26,29 | N/A | N/A | 26,57 |
| 64 | 0 | N/A | N/A | 24,92 | N/A | N/A | 25,46 |
| 65 | 0 | N/A | N/A | 26,85 | N/A | N/A | 27,61 |
| 66 | 0 | N/A | N/A | 23,13 | N/A | N/A | 24,68 |
| 67 | 0 | N/A | N/A | 26,18 | N/A | N/A | 26,86 |
| 68 | 0 | N/A | N/A | 24,44 | N/A | N/A | 25,32 |
| 69 | 0 | N/A | N/A | 23,43 | N/A | N/A | 23,54 |
| 70 | 0 | N/A | N/A | 24,36 | N/A | N/A | 24,62 |

Specificity for the triplex assay was 100% compared to the singleplex assay. Considering the triplex assay as a first screening for an extra singleplex evaluation of samples showing amplification for only N1 or N2, the sensitivity of our triplex assay protocol was 98% (Table 1). The viral loads detailed on Table 2 were calculated running a calibration curve with 2019-nCoV N positive control (IDT, USA), with a detection limit below 1 RNA copy. The viral load for the only sample that was singleplex positive but multiplex negative was 1.56 copies/uL; for the only sample singleplex assay positive but only N1 positive at triplex assay, the viral load was 33.4 copies/uL; for the two samples singleplex assay positive but only N2 positive at triplex assay, the viral loads were 22.4 and 10.3 copies/uL.

## Discussion

We herein describe the development of a triplex SARS-CoV-2 RT-qPCR for the same targets that the EUA FDA approved CDC singleplex assay. Primers and probes for triplex assay were developed by IDT (USA) which is among the few companies endorsed by CDC to purchase its CDC designed SARS-CoV-2 singleplex kit. Both the specificity and sensibility of our SARS-CoV-2 triplex RT-qPCR were 100% for a limit of detection of around 10 viral copies/uL, indicating a great performance compared with commercial tripex assays. Although the negative sample size was relatively small on this study, the specificity of N1 and N2 probes has already been tested in terms of cross reactivity with other respiratory viruses (2,4,5,6,7). Additionally, it is important to notice that this triplex SARS-CoV-2 RT-qPCR has been validated only for the instruments and chemistries we describe here and could need extra validation before implementation for others.

Although other triplex assays protocols have been recently published using some of the CDC designed primers and probes (4,5), our triplex assay protocol is the first one using the exactly same set of primers and probes than the CDC FDA EUA singleplex protocol. This CDC singleplex based triplex assay RT-qPCR represents an affordable alternative to other commercial triplex assay. For laboratories currently using the CDC protocol, this triplex assay would also speed up diagnosis while saving reagents, both necessary to improve testing capacity for SARS-CoV2.

## Data Availability

All relevant data is included in the manuscript

## Ethical considerations

All samples have been submitted for routine patient care and diagnostics at Galapagos Islands. This study was authorized by “Comité de Operaciones Especiales Regional de Galápagos” that is leading board for Covid19 surveillance in Galapagos Islands. All data used in the current study was anonymized prior to being obtained by the authors.

## Acknowledgements

We thank the medical personnel from “Ministerio de Salud Pública” at Galapagos Islands and the staff from the “Agencia de Regulación y Control de la Bioseguridad y Cuarentena para Galápagos” for their support. We also thank Dr. Ronald Cedeño from OPS/WHO for his work during Covid 19 surveillance in Galapagos Islands. We specially thank Gabriel Iturralde, Oscar Espinosa and Dr Tannya Lozada from “Dirección General de Investigación de la Universidad de Las Américas”, and also the authorities from Universidad de Las Américas, for logistic support to make SARS-CoV-2 diagnosis possible in Galapagos Islands.

## References

1. Interim Guidelines for Collecting, Handling, and Testing Clinical Specimens from Persons for Coronavirus Disease 2019 (COVID-19). Center for Diseases Control and Prevention, USA. https://www.cdc.gov/coronavirus/2019-ncov/lab/guidelines-clinical-specimens.html (last accession date 04/20/20).

2. Xiaoyan Lu, Lijuan Wang, Senthilkumar K. Sakthivel, Brett Whitaker, Janna Murray, Shifaq Kamili, Brian Lynch, Lakshmi Malapati, Stephen A. Burke, Jennifer Harcourt, Azaibi Tamin, Natalie J. Thornburg, Julie M. Villanueva and Stephen Lindstrom. US CDC Real-Time Reverse Transcription PCR Panel for Detection of Severe Acute Respiratory Syndrome Coronavirus 2. Emerging Infectious Diseases. 2020. Volume 26:8.

3. https://www.fda.gov/medical-devices/emergency-situations-medical-devices/emergency-use-authorizations (last accession date 06/16/2020).

4. Jesse J. Waggoner, Victoria Stittleburg, Renee Pond, Youssef Saklawi, Malaya K. Sahoo, Ahmed Babiker, Laila Hussaini, Colleen S. Kraft, Benjamin A. Pinsky, Evan J. Anderson, and Nadine Rouphael. Triplex Real-Time RT-PCR for Severe Acute Respiratory Syndrome Coronavirus 2. Emerging Infectious Diseases. 2020. Volume 26:7.

5. Garrett A. Perchetti, Arun K. Nalla, Meei-Li Huang, Keith R. Jerome, Alexander L. Greninger. Multiplexing primer/probe sets for detection of SARS-CoV-2 by qRT-PCR. Journal of Clinical Virology. 2020. 129:104499.

6. Daniel D. Rhoads, Sree S. Cherian, Katharine Roman, Lisa M. Stempak, Christine L. Schmotzer and Navid Sadri. Comparison of Abbott ID Now, Diasorin Simplexa, and CDC FDA EUA methods for the detection of SARS-CoV-2 from nasopharyngeal and nasal swabs from individuals diagnosed with COVID-19. Accepted Manuscript Posted Online 17 April 2020. J. Clin. Microbiol. doi:10.1128/JCM.00760-20.

7. Arun K. Nalla, Amanda M. Casto, Meei-Li W. Huang, Garrett A. Perchetti, Reigran Sampoleo, Lasata Shrestha, Yulun Wei, Haiying Zhu, Keith R. Jerome and Alexander L. Greninger. Comparative Performance of SARS-CoV-2 Detection Assays using Seven Different Primer/Probe Sets and One Assay Kit. JCM Accepted Manuscript Posted Online 8 April 2020. J. Clin. Microbiol. doi:10.1128/JCM.00557-20.

